# Time to registry discontinuity in Tanzania’s national HIV care registry: a survival analysis of population mobility patterns

**DOI:** 10.64898/2026.03.07.26347830

**Authors:** James Mwakyomo, Raphael Z. Sangeda, Hillary Mushi, Prosper Njau

## Abstract

**Background:** Early loss to follow-up (LTFU) after HIV enrolment is widely used to monitor program performance and progress toward treatment targets. These indicators assume that absence from the registering clinic reflects disengagement from care. However, in settings with substantial internal migration, patients may continue treatment at another facility while appearing to be lost in routine records. We evaluated the timing and geographic patterns of registry discontinuity following HIV registration in Tanzania to assess whether early LTFU primarily reflects patient disengagement from care or the characteristics of the monitoring system.

**Methods:** We conducted a national registry-based observational analysis using routinely collected data from the HIV care registry maintained by the National AIDS and Sexually Transmitted Infections Control Programme (NASHCoP). The analysis included 2,136,207 individuals with recorded district registration and visit dates between 2017 and 2021. Registry discontinuity was defined as the interval between the first recorded visit and the absence of further recorded visits within the registration facility. Kaplan-Meier methods were used to estimate time-to-discontinuity patterns, and persistence curves were compared across predefined population mobility corridors (stable districts, urban migration districts, mining areas, pastoralist regions, and border districts). Threshold summaries were calculated at 30, 60, 90, and 180 days.

**Results:** The median duration between the first and last recorded visits was 777 days (IQR, 217-1659). Registry discontinuity occurred predominantly soon after registration: 9.6% of individuals had no further recorded visits within 30 days, 11.8% within 60 days, 13.5% within 90 days, and 17.8% within 180 days of registration. A Kaplan-Meier analysis showed a steep early decline, followed by a prolonged plateau, indicating that most discontinuities occurred shortly after the first recorded visit. The time to registry discontinuity differed significantly across mobility corridors (log-rank p < 0.001), with earlier discontinuities in border, urban-migration, and mining districts compared with stable districts. Nearly one million individuals were recorded as newly registered in 2017, suggesting that first registry appearance frequently reflects administrative enrolment rather than first lifetime initiation of HIV care.

**Conclusions:** Early registry discontinuity following HIV registration in Tanzania is common, occurring soon after the first recorded visit, and shows a consistent geographic structure associated with population mobility. These findings indicate that a substantial proportion of apparent early LTFU reflects administrative discontinuity rather than confirmed treatment interruption. Facility-based retention indicators may therefore underestimate treatment continuity among mobile populations. Monitoring systems capable of linking patient records across facilities and administrative boundaries are required to distinguish between geographic relocation and disengagement from care.

## Introduction

The scale-up of antiretroviral therapy (ART) programs across sub-Saharan Africa has led to an increasing reliance on routine program indicators to monitor performance and progress toward international HIV treatment targets. Among these indicators, retention in care and early loss to follow-up (LTFU) after enrolment are widely used to assess program performance. Early disengagement from care has been associated with delayed treatment initiation, treatment interruption, virologic failure, and increased mortality. Therefore, programmes routinely monitor the proportion of patients who do not return after registration or an initial clinic visit. Longitudinal cohort studies from East Africa have demonstrated substantial heterogeneity in observed retention patterns despite similar treatment availability [1–3].

Early LTFU is commonly defined as the absence of a clinic visit within a specified period after enrolment, typically within 30, 60, or 90 days. In routine monitoring systems, these indicators are often interpreted as reflecting patient behavior and the quality of service delivery. High levels of early LTFU are therefore frequently attributed to barriers such as stigma, transport costs, and limited access to health facilities [4,5]. However, such interpretations rely on the implicit assumption that patients remain geographically stable and that the facility where they initially registered continues to serve as their primary care facility.

This assumption may not hold in many low- and middle-income countries. Routine HIV monitoring systems frequently track individuals only within the facilities or administrative units where they are registered. Patients who continue treatment at another facility may therefore appear indistinguishable from those who have disengaged from care, a phenomenon often described as "silent transfer" [6]. Silent transfers have been documented at both the clinic and program levels and may lead to substantial underestimation of retention in care when patient movement between facilities is not captured [7–9]. Consequently, routine program indicators may reflect the characteristics of monitoring systems and administrative recording processes rather than true patient outcomes, highlighting the need for person-centered longitudinal data systems capable of tracking individuals across facilities and regions [10].

Population mobility represents an additional challenge for interpreting retention indicators. Internal migration associated with pastoralism, seasonal agricultural work, mining employment, cross-border trade, and rural-to-urban migration is common in many parts of sub-Saharan Africa. Recent studies have increasingly recognized mobility as an important determinant of HIV care continuity and retention patterns [11,12]. When monitoring systems do not link patient records across administrative boundaries, individuals who relocate after registration may be recorded as early LTFU, even when they continue treatment elsewhere.

Tanzania provides a useful setting to examine these dynamics. The national HIV program maintains a centralized registry that records patient registration and follow-up within administrative districts but does not routinely link encounters across district boundaries. At the same time, internal population mobility is substantial. It occurs through several well-recognized patterns, including pastoral migration in northern regions, seasonal agricultural labor movements, mining-related migration, border trade corridors, and rural-to-urban movements toward major cities. In such settings, geographic relocation may result in apparent early LTFU within the registering district, even when patients continue to receive treatment elsewhere.

Recent analyses of routine programme data in Tanzania have suggested that patterns of apparent LTFU may vary substantially across ecological settings. For example, district-level studies have reported differing dynamics between long-established treatment programs in relatively stable populations and regions characterized by seasonal or labor-related mobility. Complementary analyses of pharmacy refill adherence and treatment outcomes in Tanzanian program data have similarly demonstrated strong geographic variation in retention and treatment continuity [13,14]. These observations raise the possibility that a portion of the early LTFU signals observed in national program data may reflect geographic mobility and registry structure rather than uniform patient disengagement from care.

If early LTFU partly reflects geographic relocation rather than treatment discontinuation, routine retention indicators may systematically misclassify program performance, particularly in highly mobile populations. Approaches such as unique patient identifiers, case-based surveillance systems, and biometric monitoring have been proposed to address this limitation, and studies using such systems have demonstrated that many individuals classified as lost to follow-up can later be identified as being in care elsewhere [15,16]. However, direct patient-level linkage across districts is not yet available in routine Tanzanian registry data.

In this study, we used the national HIV care registry to evaluate the timing and geographic distribution of registry discontinuity following HIV registration in Tanzania. Rather than estimating individual clinical outcomes, the objective was to examine the measurement behavior of the early LTFU indicator and assess whether its national patterns were more consistent with patient disengagement from care or administrative discontinuity associated with population mobility.

## Methods

### Study design

We conducted a national registry-based observational analysis using routinely collected HIV programme data to examine patterns of early disappearance recorded after HIV care registration. The primary objective was not to estimate clinical outcomes or identify individual predictors of LTFU, but to evaluate how an operational monitoring indicator of early LTFU behaves within a large routine health information system and whether its distribution exhibits a systematic geographic structure.

Because registry records include timestamps for the first and last recorded clinic visits within the observation window, the analysis was structured as a time-to-event evaluation of registry persistence after registration. In this context, the event of interest represents the absence of further recorded visits within the registering district rather than confirmed disengagement from HIV treatment.

### Data source

Data were obtained from an extract of the HIV care registry maintained by the National AIDS and Sexually Transmitted Infections Control Programme (NASHCoP), the national operational monitoring system for public-sector HIV services. The extract represented a programme snapshot containing registrations recorded between 1^st^ January 2017 and 31^st^ December 2021 and included approximately 2.1 million individuals.

For each individual, the dataset contained a unique programme identifier, registration region and district, first recorded clinic visit date, last recorded clinic visit date, and visit status at the time of extraction. Individuals were assigned to the administrative district corresponding to the facility responsible for care at the time of extraction within the registry.

Linked pharmacy dispensing records, laboratory monitoring results, and confirmed transfer documentation across facilities were not included in the analysis. This exclusion was intentional. The objective of this study was to evaluate how the early disappearance indicator behaves within the registry itself before correction through record linkage. Consequently, the analysis assessed how the monitoring system classifies patient engagement within districts under routine operational conditions rather than reconstructing individual clinical retention trajectories.

Because the dataset represents a time-restricted registry extract, the first recorded visit corresponds to the first visit observed within the extract window. However, it may not reflect an individual’s first lifetime entry into HIV care.

### Study population

All individuals with valid entries for district, first recorded visit date, and last recorded visit date were included in this study. Records were excluded if the dates were missing or implausible, including instances in which the last recorded visit preceded the first recorded visit.

The observation window included individuals with their first recorded visit between 1^st^ January 2017 and 31^st^ December 2021.

### Outcome definition

For each individual, the duration of registry persistence was calculated as the number of days between the first and last recorded visits within the registered district.

Early disappearance was defined as the short period between the first and last recorded visits. To characterize the timing of disappearance after registration, thresholds at 30, 60, 90, and 180 days were evaluated.

The 90-day threshold was designated as the primary spatial indicator because it approximates the period encompassing treatment initiation, stabilization, and early refill cycles within routine ART programmes. Early disappearance was therefore interpreted as the absence of further recorded visits within the registering district and was not assumed to represent confirmed treatment discontinuation.

### Handling of end-of-registry censoring

The registry extract ended on 31^st^ December 2021. Individuals registering near the end of the observation period could not accumulate sufficient follow-up time to reach the later disappearance thresholds. Including such individuals in the denominator would artificially lower the disappearance estimates.

To address this issue, threshold-specific eligibility cohorts were constructed. For each disappearance threshold, individuals were included only if their registration date allowed for at least that duration of potential observation before the end of the extraction window. For example, the 180-day analysis included only individuals registering on or before 4 July 2021.

This approach ensured that all individuals included in each analysis had equal opportunities to meet the definition of disappearance.

### Geographic classification

The analyses were conducted at both district and regional administrative levels, with districts serving as the primary spatial unit.

To evaluate structured spatial patterns, districts were grouped into population mobility corridors that reflected known movement systems in Tanzania. These included pastoralist regions in the northern rangelands, mining areas in the Lake Zone, western border regions characterized by cross-border movement, and peri-urban migration zones surrounding Dar es Salaam. The remaining districts served as a comparison group, representing relatively stable populations.

This classification was used to interpret potential geographic structure in registry disappearance patterns, rather than to establish causal relationships.

### Statistical analysis

All analyses were conducted using R (version 4.5.2).

National disappearance curves were generated by calculating the cumulative proportion of individuals whose registry persistence duration was shorter than each of the thresholds (30, 60, 90, and 180 days). The annual counts of first recorded registrations were summarized to describe the temporal patterns in registry enrolment.

At the district and regional levels, the disappearance proportions were calculated as the number of individuals meeting the threshold definition divided by the number eligible for that threshold. Spatial mapping was used to evaluate geographic clustering.

To examine differences across mobility contexts, registry persistence time was analyzed as a time-to-event outcome. The follow-up time was administratively censored 180 days after registration. Kaplan-Meier curves were constructed for each mobility corridor and compared using the log-rank test.

### Interpretation framework

The analysis evaluated whether early disappearance recorded in the registry was more consistent with individual disengagement from care or geographically structured administrative discontinuity within the monitoring system.

Evidence supporting a registry-structured phenomenon was inferred when high national disappearance proportions, spatial clustering, and systematic differences across mobility corridors occurred simultaneously and were accompanied by divergent persistence trajectories over time.

### Ethics

This study used de-identified secondary programme data. No patient contact occurred and individual identities were not accessible to the investigators. The analysis was conducted with approval from the Muhimbili University of Health and Allied Sciences Research Ethics Committee (Reference: MUHAS-REC-DA.25/111/01/28 Jan 2021) in accordance with national procedures governing the analysis of routine health programme data.

## Results

### Cohort description

The analysis included 2,136,207 individuals with valid registration and follow-up dates recorded in the national HIV care registry between 2017 and 2021. The median duration between the first and last recorded visits within the registering facility was 777 days (interquartile range, 217-1659 days).

### Early registry discontinuity after registration

To account for incomplete observations near the end of the registry period, eligibility for follow-up was defined separately for each time threshold. Using these threshold-specific eligible cohorts, the cumulative proportion of individuals without a subsequent recorded visit to the registering facility was 9.6% at 30 days, 11.8% at 60 days, 13.5% at 90 days, and 17.8% at 180 days (Table 1).

**Table 1:**
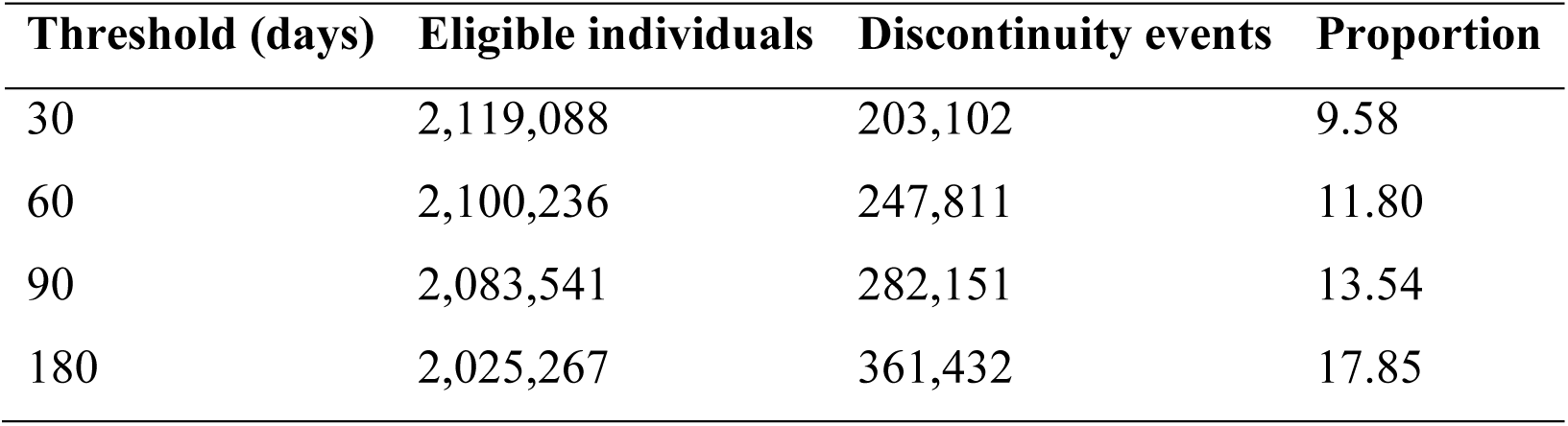
Number of individuals eligible for follow-up and proportion without a subsequent recorded visit within 30, 60, 90, and 180 days after the first recorded HIV clinic visit. Eligibility was defined separately for each threshold to account for right censoring near the end of the registry observation period (2017-2021).

| **Threshold (days)** | **Eligible individuals** | **Discontinuity events** | **Proportion** |
| --- | --- | --- | --- |
| 30 | 2,119,088 | 203,102 | 9.58 |
| 60 | 2,100,236 | 247,811 | 11.80 |
| 90 | 2,083,541 | 282,151 | 13.54 |
| 180 | 2,025,267 | 361,432 | 17.85 |

The timing of registry discontinuity following the first recorded visit showed a steep early decline, followed by a prolonged plateau, indicating that most events occurred soon after registration rather than accumulating gradually over extended follow-up. The 30-, 60-, 90-, and 180-day thresholds summarize the early portion of the time-to-discontinuity distribution (Figure 1).

**Figure 1:**
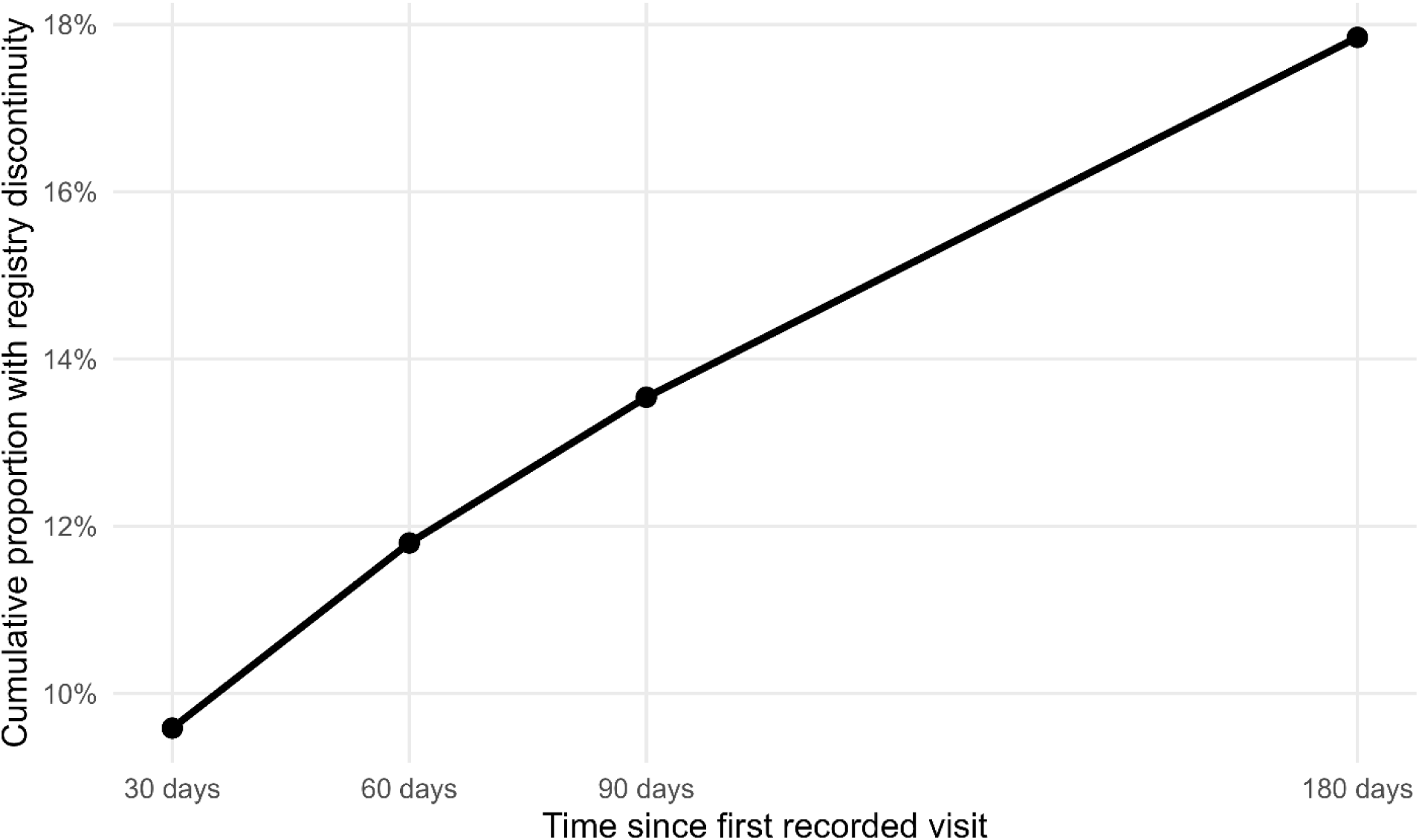
Early registry discontinuity following HIV registration. Cumulative proportion of individuals without a subsequent recorded visit to the registering facility within 30, 60, 90, and 180 days of the first recorded HIV clinic visit in the national HIV care registry. Each point represents the proportion calculated from a threshold-specific eligible cohort, accounting for correct censoring near the end of the registry observation period (2017-2021 extract).

Annual counts of the first recorded registrations varied markedly over time, with a pronounced peak in 2017, followed by lower, relatively stable levels in subsequent years (Figure 2). The large initial peak indicates that the first registry appearance often reflects administrative enrolment or database integration rather than the true first initiation of HIV care.

**Figure 2:**
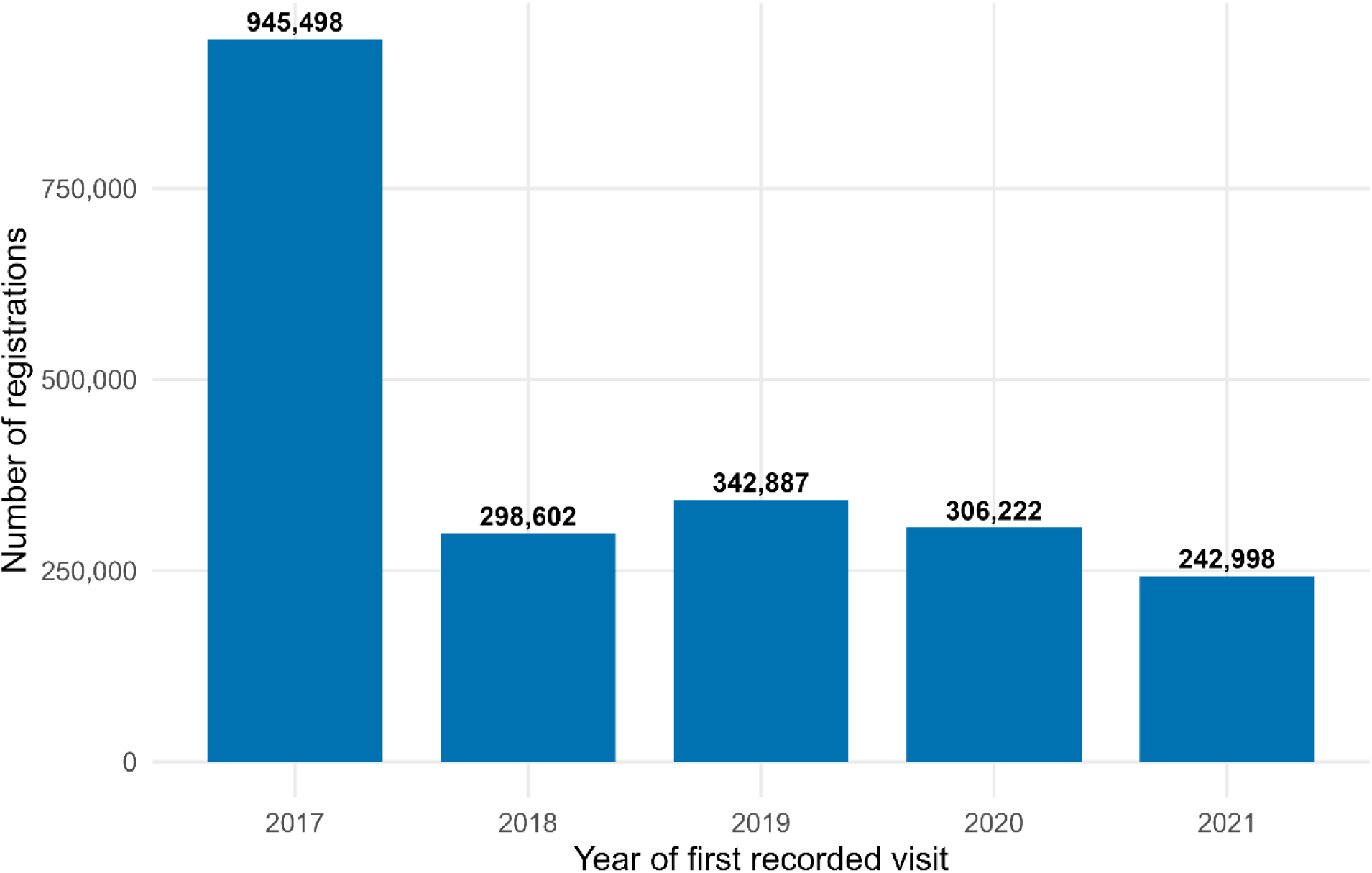
Annual first recorded registrations in the national HIV registry. Number of individuals with a first recorded HIV clinic visit in the national registry by calendar year (2017-2021). Counts represent the first appearance of an individual in the registry extract and do not necessarily correspond to the first lifetime entry into HIV care.

### Time-to-registry discontinuity

The Kaplan-Meier analysis showed that the discontinuity in the registry was concentrated shortly after registration (Figure 3). The survival curve demonstrated a steep early decline, followed by a prolonged plateau, indicating that most discontinuity events occurred soon after the first recorded visit, with a comparatively small number of additional events accumulating over time.

**Figure 3:**
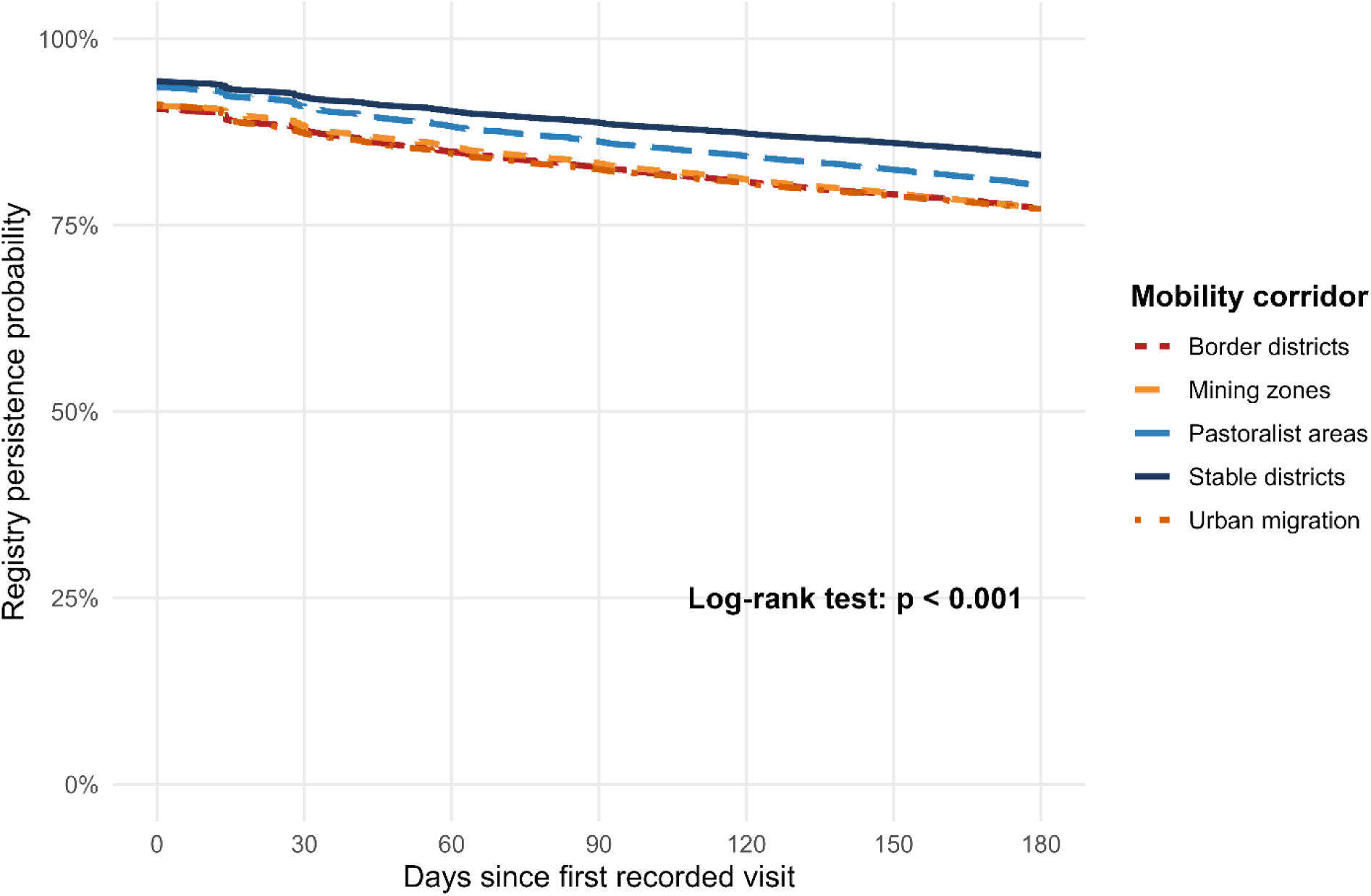
Time to registry discontinuity during the first 180 days after registration. Kaplan-Meier estimates of retention at the registering facility during the first 180 days following the first recorded HIV clinic visit, stratified by population mobility corridor. The event was defined as the absence of a subsequent recorded visit at the registering facility within 180 days. Individuals with at least 180 days of observed follow-up were censored at 180 days.

### Survival differences by population mobility corridor

Significant differences in the time to registry discontinuity were observed across population mobility contexts. Individuals registered in districts characterized by population movement showed a faster decline in observed registry persistence than those registered in relatively stable districts (Figure 3).

The Kaplan-Meier curves separated early after registration, indicating earlier registry discontinuity in mobility-associated districts. The log-rank test confirmed strong heterogeneity between mobility corridors (χ² = 15,372, df = 4, p < 0.001) (Table 2).

**Table 2:** Log-rank comparison of time-to-registry discontinuity by population mobility corridor. Results of the log-rank test comparing Kaplan-Meier time-to-registry discontinuity curves across population mobility corridors during the first 180 days after the first recorded HIV clinic visit.

| Corridor | N | Observed events | Expected events |
| --- | --- | --- | --- |
| Stable districts | 1,360,154 | 211,937 | 245,738 |
| Urban migration | 344,095 | 78,466 | 59,612 |
| Mining zones | 225,423 | 51,290 | 39,233 |
| Pastoralist areas | 63,162 | 12,381 | 11,223 |
| Border districts | 32,433 | 7,358 | 5,626 |

Log-rank $\chi^2 = 15,372$ , $df = 4$ , $p < 0.001$

Earlier RD was observed in mobility-associated districts than in those registered in stable districts. The earliest declines occurred in border districts, followed by urban migration and mining districts, whereas pastoralist districts showed intermediate patterns (Figure 3).

### Corridor-specific threshold summaries

Threshold-specific estimates also showed consistent differences in registry discontinuity across population mobility corridors. At 30 days, the proportion without further recorded visits was 13.6% in border districts, 13.4% in urban migration districts, 12.6% in mining districts, 10.4% in pastoralist districts, and 9.1% in stable districts.

After 90 days, the corresponding proportions were 20.1%, 19.5%, 19.2%, 16.6%, and 14.0%, respectively (Figure 4; Supplementary Table S1). The same ordering persisted at 180 days (27.4%, 26.1%, 27.2%, 24.1%, and 20.0%).

**Figure 4:**
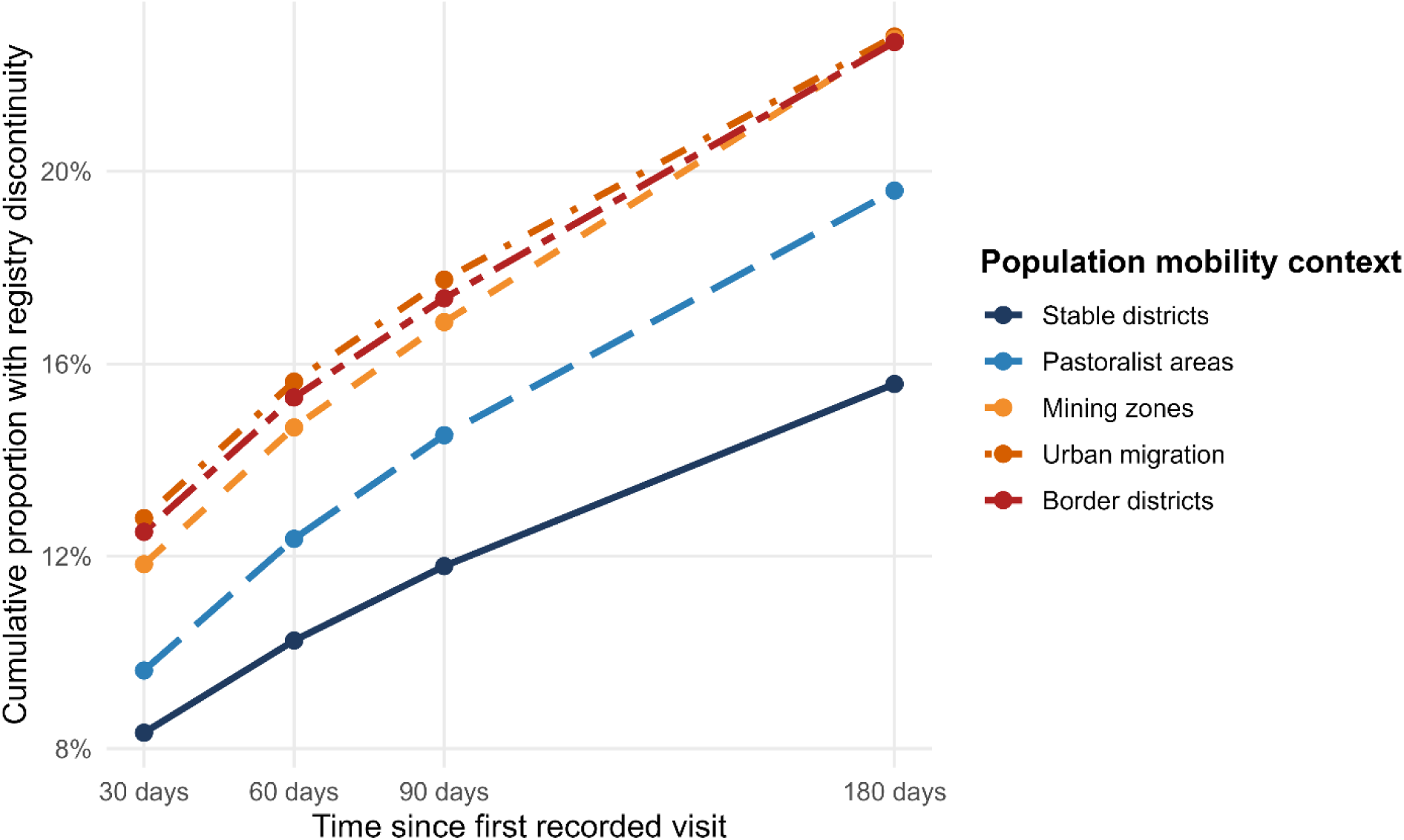
Registry discontinuity by population mobility corridor. The cumulative proportion of individuals without additional recorded visits within 30, 60, 90, and 180 days after the first recorded HIV clinic visit, stratified by population mobility context (stable districts, urban migration districts, mining zones, pastoralist areas, and border districts). Districts associated with population mobility contexts consistently showed earlier termination of the observed follow-up than stable districts.

### Geographic heterogeneity

Substantial geographic variation in the observed registry discontinuity was observed across Tanzania (Figure 5).

**Figure 5:**
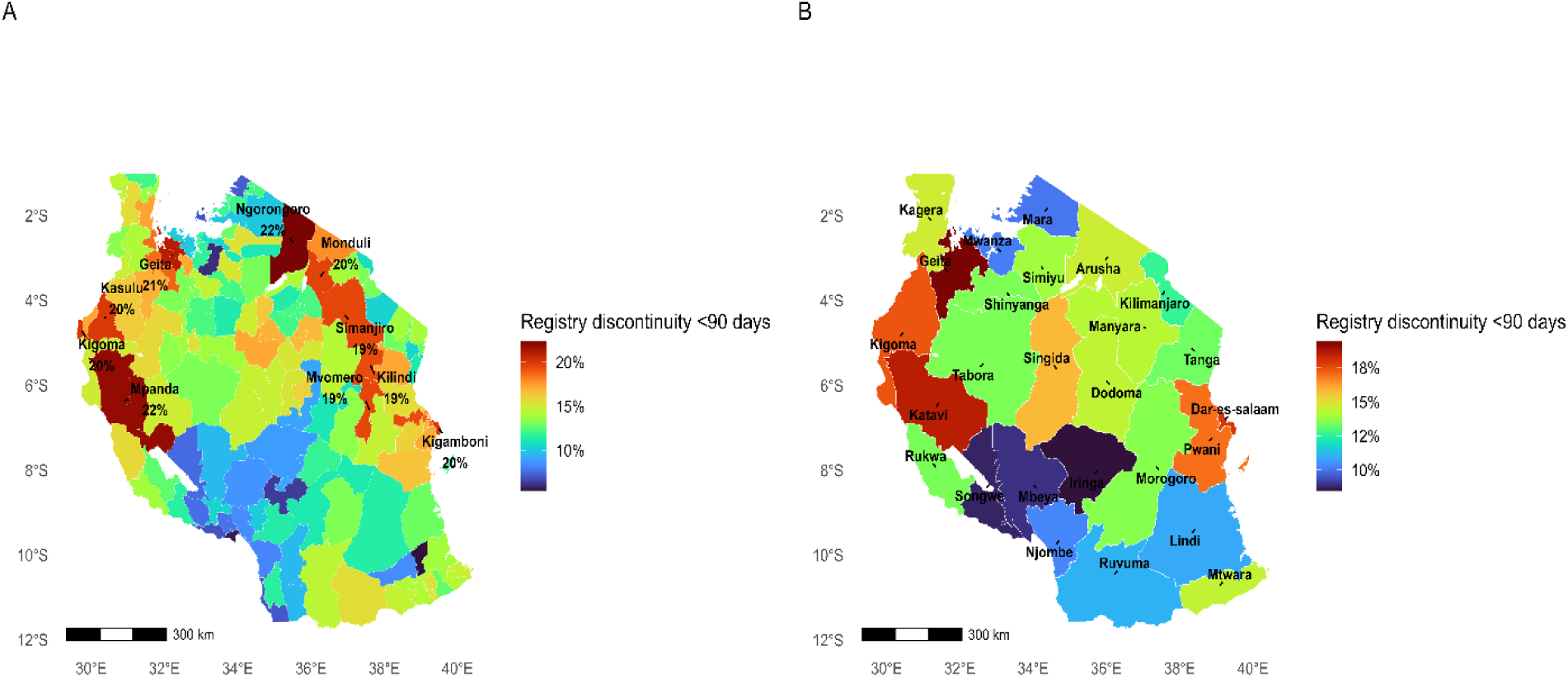
Geographic distribution of early registry discontinuity (90-day) Panel A shows the district-level proportion of individuals who did not have a subsequent recorded visit within 90 days of the first recorded HIV clinic visit. Panel B shows the regional-level proportions using the same eligibility criteria. Estimates were calculated using threshold-specific eligible cohorts to account for right censoring near the end of the registry observation period (2017-2021 extraction).

Only individuals eligible for a 90-day follow-up before December 31, 2021, were included. Zanzibar was excluded because of differences in registry coverage.

At the district level, the highest 90-day discontinuity proportions were observed in the Ngorongoro (24.4%), Mpanda (24.3%), Kilindi (22.9%), Kasulu (22.7%), and Mbogwe (22.4%) districts (Supplementary Table S2). Several adjacent districts exceeded 20%.

At the regional level, the highest 90-day discontinuity proportions were observed in Katavi (21.2%), Geita (21.1%), and Kigoma (20.1%). Intermediate levels were observed in Dar es Salaam (17.6%) and Pwani (17.0%), whereas the lowest values were observed in Iringa (9.6%), Songwe (11.1%), Mbeya (11.2%), and Njombe (11.8%) (Supplementary Table S3).

Neighboring districts and regions tended to show similar levels of registry discontinuity.

## Discussion

The early disappearance thresholds examined in this study correspond to the initial portion of the registry persistence curve and therefore capture events occurring shortly after registration rather than long-term attrition. A Kaplan-Meier analysis demonstrated a pronounced early decline, followed by a prolonged plateau, indicating that most registry disappearances occurred soon after the first recorded visit, with substantially lower rates thereafter. This front-loaded pattern differs from the gradual behavioral disengagement typically observed in longitudinal treatment cohorts [1–3,17] and instead suggests early separation of administrative registry records.

At the national level, early registry disappearance was common and occurred predominantly within the first three months after the first recorded visit. These events showed a consistent geographic structure across districts and regions. Earlier discontinuity occurred in districts characterized by population mobility, including pastoralist regions, mining areas, border districts, and peri-urban migration zones. Similar associations between population mobility and observed retention patterns have been reported in several sub-Saharan African settings [11,12]. The concentration of early registry disappearance in mobility-associated districts suggests that geographic movement may play an important role in shaping routine monitoring indicators.

The large number of individuals recorded as newly registered during the first year of observation provides additional evidence that registry appearance does not necessarily correspond to the first lifetime initiation of HIV care. Instead, the first recorded visit in the registry often reflects administrative enrolment within a district or the integration of previously existing patient records into the national system. Similar observations have been reported in studies using laboratory histories and record-linkage approaches, in which individuals classified as newly enrolled were found to have evidence of prior treatment elsewhere [9].

Taken together, these findings indicate that early LTFU signals recorded in routine program data do not solely measure patient retention. A substantial component may reflect the structure of monitoring systems themselves, particularly when patient records cannot be linked across facilities or administrative regions. When individuals relocate and subsequently enroll at another facility, they may be recorded as lost in one district while continuing treatment in another. Such "silent transfers" have been documented in several HIV treatment programs and surveillance systems [6–8].

The structure of routine registry systems can amplify this phenomenon. In the monitoring system analyzed here, each patient is linked to a single facility or administrative district at a given time. When relocation occurs, a new record may be created at the receiving facility without linkage to the prior record. The original facility records a rapid loss, whereas the receiving facility records new enrolment. This process simultaneously increases the number of apparent new registrations and inflates early LTFU indicators.

These findings have important implications for the interpretation of routine HIV program indicators. Facility-level retention measures may underestimate program performance in highly mobile populations and exaggerate geographic differences in retention. Districts characterized by seasonal or economic mobility may appear to perform poorly, even when individuals remain in treatment elsewhere. Systems capable of linking patient records across facilities would allow programs to measure retention in care rather than persistence within a single facility registry. Person-centered surveillance systems and unique patient identifiers have been proposed to address this limitation [10,15] and biometric monitoring systems have demonstrated that individuals recorded as lost are often subsequently identified in care elsewhere [16].

These findings complement other analyses of Tanzanian HIV programme data, demonstrating geographic variation in treatment continuity and retention indicators. Studies examining geographic variation in LTFU and pharmacy refill adherence have reported substantial regional differences in treatment continuity within routine programme data [13,14]. District-level investigations in Tanzania have also documented contrasting retention dynamics between long-established treatment programmes in relatively stable populations and regions characterized by seasonal or labor-related mobility [18,19]. Together, these observations suggest that retention patterns observed in routine programme indicators may vary considerably across geographic contexts, underscoring the importance of interpreting early LTFU signals within the spatial and programmatic structure of the monitoring system.

This study has several limitations inherent to administrative registry data. The registry records the district responsible for care at registration, but does not capture subsequent encounters across facilities. Transfers between districts, therefore, could not be directly observed, and the destination of care following the registry’s disappearance was unknown. Early disappearance was defined as the absence of further recorded visits within the registering district rather than confirmed treatment cessation. Consequently, the analysis could not distinguish true disengagement from undocumented transfers or re-registration elsewhere. In addition, mobility was inferred from spatial and temporal patterns rather than measured directly, a common approach when individual movement data are unavailable [11].

In conclusion, early disappearance recorded in the national HIV care registry after registration in Tanzania is common, occurs predominantly soon after the first recorded visit, and shows a consistent geographic structure. The timing and concentration of events in districts characterized by population mobility suggest that a substantial proportion of early LTFU reflects administrative discontinuity associated with patient movement across districts rather than confirmed treatment interruption. Monitoring systems capable of linking patient records across administrative boundaries will be essential for accurately measuring retention in care and designing appropriate program responses in highly mobile populations.

## Declarations

### Ethics approval and consent to participate

This study used de-identified secondary data obtained from the national HIV care registry maintained by the National AIDS and Sexually Transmitted Infections Control Programme (NASHCoP). No direct patient contact occurred and individual identities were not accessible to the investigators. The analysis was approved by the Muhimbili University of Health and Allied Sciences Research Ethics Committee (MUHAS-REC) (Reference: MUHAS-REC-DA.25/111/01/28 January 2021) in accordance with national procedures governing the use of routine health program data.

### Availability of data and materials

The data used in this study were derived from the national HIV care registry maintained by the National AIDS and Sexually Transmitted Infections Control Programme (NASHCoP). They are not publicly available due to programme data governance and confidentiality restrictions. Access to the data may be requested from NASHCoP, subject to institutional approval.

## Acknowledgements

The authors thank the National AIDS and Sexually Transmitted Infections Control Programme (NASHCoP) for maintaining the national HIV care registry and for facilitating access to routine programme monitoring data used in this analysis.

