## Supplementary Materials for "Time to registry discontinuity in Tanzania’s national HIV care registry: a survival analysis of population mobility patterns"

### SUPPLEMENTARY MATERIAL

#### Supplementary Table S1: Early registry discontinuity by population mobility corridor

Proportion of individuals without a subsequent recorded visit within 30, 60, 90, and 180 days after the first recorded HIV clinic visit, stratified by population mobility corridor. Estimates were calculated using threshold-specific eligible cohorts to minimize right censoring near the end of the observation period (2017–2021 registry extract).

| **Mobility corridor** | **30 days (%)** | **60 days (%)** | **90 days (%)** | **180 days (%)** |
|---|---|---|---|---|
| Border districts | 13.6 | 17.4 | 20.1 | 27.4 |
| Urban migration districts | 13.4 | 16.8 | 19.5 | 26.1 |
| Mining districts | 12.6 | 16.2 | 19.2 | 27.2 |
| Pastoralist districts | 10.4 | 13.9 | 16.6 | 24.1 |
| Stable districts | 9.1 | 11.8 | 14.0 | 20.0 |

**Supplementary Table S2: District-level registry discontinuity within 30, 60, 90, and 180 days after the first recorded HIV clinic visit (sorted by 90-day proportion)**

Proportion of individuals without a subsequent recorded visit at the registering facility within each threshold period. Estimates were calculated using threshold-specific eligible cohorts to minimize right censoring near the end of the registry observation period (2017–2021 extract).

| **Region** | **District** | **30 days (%)** | **60 days (%)** | **90 days (%)** | **180 days (%)** |
|---|---|---|---|---|---|
| Arusha | Ngorongoro | 15.0 | 18.8 | 24.4 | 35.7 |
| Katavi | Mpanda | 15.2 | 19.6 | 24.3 | 35.0 |
| Tanga | Kilindi | 14.8 | 18.9 | 22.9 | 32.6 |
| Kigoma | Kasulu | 14.7 | 18.7 | 22.7 | 32.6 |
| Geita | Mbogwe | 14.0 | 18.5 | 22.4 | 31.6 |
| Geita | Bukombe | 13.6 | 17.7 | 21.7 | 30.7 |
| Geita | Nyang'hwale | 13.7 | 17.8 | 21.6 | 30.6 |
| Geita | Geita | 13.5 | 17.3 | 21.1 | 29.3 |
| Katavi | Mlele | 13.1 | 16.9 | 20.7 | 29.8 |
| Kigoma | Kakonko | 12.7 | 16.9 | 20.6 | 29.5 |
| Kigoma | Kibondo | 12.5 | 16.6 | 20.4 | 29.4 |
| Kigoma | Buhigwe | 12.3 | 16.4 | 20.2 | 29.0 |
| Dar es Salaam | Kigamboni | 12.7 | 15.8 | 20.0 | 29.7 |
| Pwani | Kibaha | 12.6 | 15.9 | 20.0 | 29.2 |
| Kigoma | Kigoma | 12.3 | 16.0 | 19.7 | 28.2 |
| Arusha | Monduli | 12.2 | 15.9 | 19.6 | 27.6 |
| Kigoma | Uvinza | 12.0 | 15.8 | 19.6 | 28.4 |
| Dar es Salaam | Temeke | 12.1 | 15.2 | 19.5 | 28.8 |
| Geita | Chato | 12.6 | 16.1 | 19.5 | 27.5 |
| Arusha | Simanjiro | 12.0 | 14.8 | 18.9 | 27.5 |
| Manyara | Simanjiro | 12.0 | 14.8 | 18.9 | 27.5 |
| Arusha | Longido | 13.0 | 15.9 | 17.9 | 24.0 |
| Singida | Singida | 11.3 | 14.2 | 17.5 | 24.5 |
| Pwani | Bagamoyo | 11.2 | 14.3 | 17.2 | 24.6 |
| Pwani | Mkuranga | 11.0 | 14.1 | 17.0 | 24.3 |
| Pwani | Kisarawe | 11.0 | 14.0 | 16.9 | 24.2 |
| Pwani | Mafia | 10.9 | 14.0 | 16.9 | 24.0 |
| Dodoma | Chamwino | 10.6 | 13.6 | 16.7 | 23.7 |
| Pwani | Rufiji | 10.7 | 13.6 | 16.4 | 23.5 |

| | | | | | |
|---|---|---|---|---|---|
| Shinyanga | Kahama | 10.3 | 13.4 | 16.3 | 23.1 |
| Dar es Salaam | Kinondoni | 11.3 | 13.9 | 16.2 | 22.4 |
| Simiyu | Bariadi | 10.3 | 13.3 | 16.1 | 22.9 |
| Singida | Iramba | 10.8 | 13.6 | 16.1 | 22.1 |
| Dar es Salaam | Ilala | 11.8 | 14.3 | 16.0 | 20.8 |
| Manyara | Kiteto | 10.4 | 13.3 | 16.0 | 22.7 |
| Simiyu | Masalala | 10.2 | 13.2 | 16.0 | 22.9 |
| Mtwara | Mtwara | 9.8 | 12.8 | 15.9 | 22.3 |
| Rukwa | Nkasi | 10.4 | 13.4 | 15.9 | 22.0 |
| Simiyu | Busega | 10.2 | 13.1 | 15.9 | 22.6 |
| Singida | Manyoni | 10.7 | 13.4 | 15.9 | 22.0 |
| Kagera | Bukoba | 10.4 | 13.4 | 15.8 | 22.2 |
| Rukwa | Sumbawanga | 10.3 | 13.2 | 15.8 | 21.8 |
| Simiyu | Itilima | 10.1 | 13.0 | 15.8 | 22.6 |
| Shinyanga | Shinyanga | 10.0 | 13.0 | 15.7 | 22.4 |
| Tabora | Tabora | 10.4 | 13.2 | 15.5 | 21.4 |
| Manyara | Babati | 10.3 | 13.1 | 15.4 | 21.4 |
| Manyara | Babati Rural | 10.2 | 13.0 | 15.4 | 21.5 |
| Dodoma | Dodoma | 10.4 | 13.0 | 15.3 | 21.1 |
| Manyara | Haneti | 10.0 | 12.8 | 15.3 | 21.8 |
| Tabora | Uyui | 10.3 | 13.1 | 15.3 | 21.2 |
| Manyara | Mbulu | 10.0 | 12.7 | 15.2 | 21.5 |
| Rukwa | Sumbawanga Rural | 10.1 | 12.9 | 15.2 | 20.9 |
| Tabora | Kaliua | 10.3 | 13.0 | 15.2 | 21.0 |
| Kagera | Bukoba Rural | 9.9 | 12.8 | 15.1 | 21.8 |
| Tabora | Igunga | 10.2 | 12.9 | 15.1 | 20.9 |
| Tabora | Urambo | 10.1 | 12.8 | 15.0 | 20.8 |
| Shinyanga | Kishapu | 9.8 | 12.5 | 14.9 | 20.9 |
| Tabora | Nzega | 10.1 | 12.8 | 14.9 | 20.6 |
| Kagera | Biharamulo | 9.3 | 12.2 | 14.8 | 21.4 |
| Rukwa | Kalama | 10.0 | 12.7 | 14.8 | 20.3 |
| Dodoma | Bahi | 10.0 | 12.5 | 14.7 | 19.9 |
| Tabora | Sikonge | 10.0 | 12.6 | 14.7 | 20.3 |
| Kagera | Uyui | 9.4 | 12.2 | 14.6 | 21.0 |
| Tanga | Handeni | 9.9 | 12.5 | 14.6 | 19.8 |
| Tanga | Tanga | 9.8 | 12.4 | 14.5 | 19.8 |
| Dodoma | Chemba | 9.2 | 11.7 | 14.4 | 20.4 |
| Morogoro | Morogoro | 9.4 | 12.1 | 14.4 | 20.3 |
| Tanga | Korogwe | 9.8 | 12.3 | 14.3 | 19.3 |
| Tanga | Mkinga | 9.8 | 12.3 | 14.3 | 19.3 |
| Morogoro | Uluguru | 9.2 | 11.9 | 14.2 | 20.1 |

| | | | | | |
|---|---|---|---|---|---|
| Tanga | Magomeni | 9.7 | 12.2 | 14.2 | 19.1 |
| Tanga | Muheza | 9.7 | 12.2 | 14.2 | 19.1 |
| Arusha | Karatu | 9.8 | 12.2 | 14.1 | 18.6 |
| Kagera | Kyerwa | 9.1 | 11.8 | 14.1 | 20.3 |
| Kagera | Muleba | 9.2 | 11.8 | 14.1 | 20.5 |
| Arusha | Arusha | 9.4 | 12.1 | 14.0 | 19.0 |
| Kagera | Ngara | 9.2 | 11.7 | 14.0 | 20.1 |
| Morogoro | Kilosa | 9.3 | 11.9 | 14.0 | 19.8 |
| Tanga | Pangani | 9.6 | 12.0 | 14.0 | 18.9 |
| Kagera | Missenyi | 9.0 | 11.5 | 13.9 | 20.0 |
| Mtwara | Masasi | 9.4 | 12.1 | 13.9 | 18.9 |
| Mwanza | Nyamagana | 9.1 | 11.8 | 13.9 | 19.3 |
| Tanga | Lushoto | 9.5 | 11.8 | 13.9 | 18.7 |
| Dodoma | Kondoa | 9.2 | 11.5 | 13.8 | 19.4 |
| Morogoro | Kilombero | 9.2 | 11.8 | 13.8 | 19.5 |
| Kagera | Karagwe | 8.7 | 11.3 | 13.7 | 19.7 |
| Morogoro | Gairo | 9.1 | 11.7 | 13.7 | 19.4 |
| Dodoma | Mpwapwa | 9.0 | 11.3 | 13.6 | 19.3 |
| Mara | Musoma | 8.9 | 11.5 | 13.6 | 18.9 |
| Morogoro | Mvomero | 9.0 | 11.6 | 13.6 | 19.2 |
| Mwanza | Ilemela | 8.9 | 11.5 | 13.5 | 18.6 |
| Lindi | Lindi | 9.0 | 11.5 | 13.4 | 18.6 |
| Mtwara | Nanyumbu | 9.2 | 11.7 | 13.4 | 18.1 |
| Mtwara | Newala | 9.2 | 11.7 | 13.3 | 18.0 |
| Kilimanjaro | Mwanga | 8.6 | 11.1 | 13.2 | 18.5 |
| Mara | Musoma Rural | 8.8 | 11.2 | 13.2 | 18.4 |
| Mtwara | Tandahimba | 9.1 | 11.6 | 13.2 | 17.8 |
| Dodoma | Kongwa | 8.7 | 10.9 | 13.1 | 18.7 |
| Mwanza | Misungwi | 8.7 | 11.2 | 13.1 | 18.1 |
| Kilimanjaro | Rombo | 8.4 | 10.8 | 13.0 | 18.3 |
| Mara | Serengeti | 8.7 | 11.2 | 13.0 | 17.8 |
| Ruvuma | Songea | 9.0 | 11.5 | 13.0 | 17.2 |
| Mara | Bunda | 8.7 | 11.1 | 12.9 | 17.6 |
| Mwanza | Kwimba | 8.6 | 11.0 | 12.9 | 17.7 |
| Kilimanjaro | Moshi | 8.2 | 10.7 | 12.8 | 18.0 |
| Lindi | Nachingwea | 8.8 | 11.3 | 12.8 | 17.4 |
| Mara | Rorya | 8.5 | 10.9 | 12.8 | 17.9 |
| Kilimanjaro | Moshi Rural | 8.1 | 10.5 | 12.7 | 17.9 |
| Lindi | Kilwa | 8.7 | 11.2 | 12.7 | 17.1 |
| Kilimanjaro | Same | 8.2 | 10.5 | 12.6 | 17.8 |
| Mwanza | Magu | 8.5 | 10.8 | 12.6 | 17.2 |
| Ruvuma | Mbinga | 8.8 | 11.1 | 12.6 | 16.7 |
| Lindi | Liwale | 8.5 | 11.0 | 12.5 | 16.9 |

| | | | | | |
|---|---|---|---|---|---|
| Ruvuma | Tunduru | 8.7 | 11.0 | 12.5 | 16.6 |
| Arusha | Arumeru | 8.1 | 10.5 | 12.4 | 15.9 |
| Ruvuma | Songea Rural | 8.7 | 11.0 | 12.4 | 16.4 |
| Kilimanjaro | Hai | 8.0 | 10.3 | 12.2 | 16.8 |
| Lindi | Ruangwa | 8.3 | 10.7 | 12.2 | 16.6 |
| Njombe | Njombe | 8.3 | 10.6 | 12.1 | 16.1 |
| Njombe | Makambako | 8.2 | 10.4 | 11.9 | 15.8 |
| Njombe | Ludewa | 8.1 | 10.3 | 11.7 | 15.5 |
| Njombe | Wanging'ombe | 8.1 | 10.3 | 11.7 | 15.5 |
| Mbeya | Chunya | 7.8 | 10.1 | 11.6 | 16.0 |
| Mbeya | Mbeya | 7.7 | 10.0 | 11.5 | 15.9 |
| Njombe | Makete | 8.0 | 10.1 | 11.5 | 15.1 |
| Songwe | Mbozi | 7.7 | 9.8 | 11.2 | 15.4 |
| Mbeya | Busokelo | 7.6 | 9.6 | 11.1 | 15.3 |
| Mbeya | Kyela | 7.7 | 9.7 | 11.1 | 15.5 |
| Songwe | Momba | 7.6 | 9.7 | 11.1 | 15.2 |
| Mbeya | Mbarali | 7.6 | 9.7 | 11.0 | 15.1 |
| Mbeya | Mbeya Rural | 7.5 | 9.6 | 11.0 | 15.3 |
| Songwe | Ileje | 7.6 | 9.6 | 11.0 | 15.1 |
| Iringa | Mafinga | 6.8 | 8.6 | 10.0 | 13.4 |
| Iringa | Iringa Rural | 6.7 | 8.5 | 9.9 | 13.4 |
| Iringa | Iringa | 6.6 | 8.4 | 9.6 | 12.6 |
| Iringa | Kilolo | 6.3 | 8.0 | 9.3 | 12.6 |

**Supplementary Table S3: Regional proportions of registry discontinuity within 30, 60, 90, and 180 days after the first recorded HIV clinic visit**

Proportion of individuals without a subsequent recorded visit at the registering facility within each threshold period at the regional level. Estimates were calculated using threshold-specific eligible cohorts to minimize right censoring near the end of the registry observation period (2017–2021 extract).

| **Region** | **30 days (%)** | **60 days (%)** | **90 days (%)** | **180 days (%)** |
|---|---|---|---|---|
| Katavi | 14.10 | 18.20 | 21.20 | 30.60 |
| Geita | 13.50 | 17.40 | 21.10 | 29.80 |

| | | | | |
|---|---|---|---|---|
| Kigoma | 12.70 | 16.70 | 20.10 | 29.10 |
| Dar es Salaam | 11.80 | 14.60 | 17.60 | 25.40 |
| Pwani | 11.10 | 14.10 | 17.00 | 24.30 |
| Singida | 10.90 | 13.80 | 16.50 | 22.90 |
| Simiyu | 10.20 | 13.20 | 16.00 | 22.80 |
| Manyara | 10.30 | 13.10 | 15.70 | 22.30 |
| Shinyanga | 10.00 | 13.00 | 15.70 | 22.40 |
| Rukwa | 10.20 | 13.10 | 15.40 | 21.30 |
| Tabora | 10.20 | 13.00 | 15.20 | 21.00 |
| Arusha | 9.90 | 12.60 | 14.80 | 20.30 |
| Tanga | 10.00 | 12.60 | 14.70 | 20.00 |
| Kagera | 9.30 | 12.00 | 14.40 | 20.60 |
| Dodoma | 9.60 | 12.10 | 14.30 | 19.90 |
| Morogoro | 9.20 | 11.80 | 14.00 | 19.80 |
| Mtwara | 9.40 | 12.00 | 13.90 | 18.90 |
| Mwanza | 8.80 | 11.30 | 13.30 | 18.40 |
| Mara | 8.70 | 11.20 | 13.10 | 18.10 |
| Kilimanjaro | 8.20 | 10.60 | 12.70 | 17.80 |
| Lindi | 8.70 | 11.10 | 12.70 | 17.30 |
| Ruvuma | 8.80 | 11.10 | 12.60 | 16.70 |
| Njombe | 8.10 | 10.30 | 11.80 | 15.60 |
| Mbeya | 7.70 | 9.80 | 11.20 | 15.50 |
| Songwe | 7.60 | 9.70 | 11.10 | 15.20 |
| Iringa | 6.60 | 8.40 | 9.60 | 12.90 |
